# A school-based randomized clinical trial of silver diamine fluoride for dental caries incidence

**DOI:** 10.1101/2023.08.16.23294171

**Authors:** Ryan Richard Ruff, Tamarinda Barry Godín, Richard Niederman

## Abstract

**Importance:** Dental caries is the world’s most prevalent noncommunicable disease and a source of severe health inequity. To prevent and reduce this burden, the Centers for Disease Control and Prevention recommends school dental sealant programs.

**Objective:** To determine whether silver diamine fluoride (SDF) is non-inferior to dental sealants and atraumatic restorative treatment (ART) for dental caries when used in a school-based program.

**Design:** The CariedAway study was a cluster-randomized, single-blind, pragmatic non-inferiority trial conducted from 2018-2023. Four years of follow-up were included.

**Setting:** Primary schools in New York City with at least 55% of the student population reporting as Black or Hispanic/Latino and at least 80% receiving free or reduced lunch.

**Participants:** Any child between the ages of 5 and 13 was eligible. There were 17741 eligible children across 48 schools.

**Interventions:** Participants were cluster-randomized at the school level to receive either a 38% concentration SDF solution or glass ionomer sealants and ART. Each participant also received fluoride varnish.

**Main Outcomes:** Primary study outcomes were the prevalence and incidence of dental caries. Our a priori hypothesis was that SDF was non-inferior to sealants and ART in reducing caries prevalence.

**Results:** A total of 7418 children were enrolled and treated, of which 4100 completed at least one follow-up observation (55%). The overall baseline prevalence of dental caries was approximately 27% (95% CI = 25.7, 28.6). Following treatment, the odds of decay prevalence decreased longitudinally (OR = 0.79, 95% CI = 0.75, 0.83) and SDF was non-inferior compared to sealants and ART (OR = 0.94, 95% CI = 0.80, 1.11). The crude incidence of dental caries in children treated with SDF was 10.2 per 1,000 tooth-years, versus 9.8 per 1,000 tooth-years in children treated with sealants and ART, for a rate ratio of 1.046 (95% CI = 0.97, 1.12).

**Conclusions and Relevance:** In a pragmatic trial, application of silver diamine fluoride resulted in nearly identical caries incidence compared to dental sealants and ART and was non-inferior in the longitudinal prevalence of caries. SDF is an effective alternative for use in school caries prevention, increasing access and reducing costs for oral healthcare.

**Trial Registration:** ClinicalTrials.gov, #NCT03442309

## Introduction

Dental caries—the “silent epidemic”—is the world’s most prevalent noncommunicable disease^1^. The National Institute of Dental and Craniofacial Research estimates that over 50% of US children between the ages of 6 and 8 have experienced caries, with some minority groups exceeding 70% ^2^. The United Nations General Assembly considers oral diseases to be a major global burden that shares common risk factors with other noncommunicable diseases, and the World Health Organization’s (WHO) Global Oral Health Action Plan names oral disease prevention as a primary strategic objective, recommending the use of cost-effective, community-based methods to prevent caries ^3^. In 2022, the WHO added glass ionomer sealants and silver diamine fluoride (SDF) to its Model List of Essential Medicines for the first time ^4^.

Despite increases in Medicaid entitlements for dental benefits, there remain persistent access challenges to oral disease prevention throughout the United States; over 69 million Americans live in dental care health professional shortage areas ^5^. The Centers for Disease Control and Prevention recommends school sealant programs to increase access to care, reduce the prevalence of caries, and improve health equity ^6^. Dental sealants can prevent the onset of carious lesions and arrests them in the early stages ^7^, and are effective in both children and adolescents ^8^. However, the burgeoning costs of care limits the utilization of school sealant programs ^9^. Alternatively, silver diamine fluoride (SDF) is used internationally as an economically efficient treatment for caries. In clinical studies, SDF application prevents caries in the primary dentition compared to placebo ^10^, is comparable to dental sealants ^10^, and arrests existing caries ^11^. Silver diamine fluoride can be applied in as little as ten seconds ^12^ and is an inexpensive strategy to reduce the burden of caries, particularly in under resourced areas ^13^. In 2017, the United States Food and Drug Administration granted breakthrough therapy status to SDF ^14^.

The CariedAway pragmatic ^15^ trial investigated the use of silver diamine fluoride as an alternative therapy for community-based caries control and prevention ^16^. Primary clinical outcomes for CariedAway included the non-inferiority of SDF compared to dental sealants and atraumatic restorative treatment (ART) in the two year arrest of dental caries ^17^ and the non-inferiority of treatment in the four-year prevalence of caries. Secondary outcomes included oral health-related quality of life ^18^, academic performance ^19^, and the effectiveness of registered nurses in the treatment of caries with SDF ^20^. Here we report on the cumulative incidence and prevalence of caries over four years.

## Methods

### Design and Participants

CariedAway was a longitudinal, cluster-randomized, single-blind, non-inferiority pragmatic clinical trial conducted from 1 February 2019 to 1 June 2023. The trial was approved by the New York University School of Medicine Institutional Review Board (#i7-00578) and is registered at www.clinicaltrials.gov (#NCT03442309). A complete trial protocol is publicly-accessible ^16^.

Participants for CariedAway were enrolled through a two-stage process. In the first stage, any school in the New York City metropolitan area with a total student population consisting of at least 50% Hispanic/Latino or black ethnicities and at least 80% receiving free or reduced lunch was eligible for inclusion. In the second stage, any child with parental informed consent and assent in participating schools was enrolled. While any child meeting these criteria was enrolled, inclusion into analysis was restricted to those aged 5-13 years. Additional exclusion criteria included if the school had a pre-existing oral health program or provider (Stage 1) or if the child did not speak English (Stage 2).

### Interventions and Procedures

Our primary experimental condition consisted of a 38% silver diamine fluoride solution (2.24 F-ion mg/dose). We selected glass ionomer cement (GIC) sealants and atraumatic restorations as our active comparator. Atraumatic restorative treatment follows the same procedure and uses the same materials as interim therapeutic restorations (ITR), but is preferred in cases where access to traditional dental care may be limited ^21^. Both SDF and ITR are included in the American Academy of Pediatric Dentistry (AAPD) Policy on Minimally Invasive Dentistry. Each participant also received a 5% Sodium Fluoride (NaF) application.

For the experimental treatment, petroleum jelly was first applied to the lips and surrounding skin to prevent temporary staining that can result from direct contact of SDF with the soft tissue. Isolation was achieved by placing gauze and cotton rolls between the teeth to be treated and the tongue and cheek. One to two drops of silver diamine fluoride were dispensed into a mixing well and applied using a micro applicator to all posterior asymptomatic cavitated lesions as well as pits and fissures of premolars and molars. The material was agitated on the surface of all cavities using a scrubbing motion for a minimum of 30 seconds, followed by 60 seconds of air drying time. One unit dose of fluoride varnish was then applied to all teeth to mask the bitter, metallic taste of SDF. The procedure was then replicated at follow-up.

For the active control, cavity conditioner was first applied to pits and fissures for 10 seconds. GIC capsules were mixed for 10 seconds at 4,000 RPM and then applied directly via the finger-sweep technique to all pits and fissures of bicuspids and molars, ensuring that closed margins were achieved. Atraumatic restorations were also placed on asymptomatic cavitated lesions, and fluoride varnish was finally applied to all teeth. At successive observations, sealants were reapplied to any unsealed or partially sealed bicuspids and molars.

Treatments in the experimental group were provided by either dental hygienists or registered nurses. Treatments in the active control were provided by dental hygienists. All treatments were provided in a dedicated room in each school using mobile equipment and under the supervision of a licensed dentist. No personalization of the treatment plan was required or performed.

### Examiners

All dental hygienists and registered nurses received identical training in September of each year, prior to the start of the academic year for study participants. Training consisted of didactic and experiential activities, including screening, treatment protocol standardization exercises, and mock patient interactions. A total of fifteen clinical staff participated in CariedAway, all of whom were registered dental hygienists or registered nurses, licensed in New York, with previous experience or interest in pediatric patient care and community health. Due to the impact of COVID-19, there was attrition amongst the clinical staff. New clinical personnel received identical training and standardization throughout the entirety of the CariedAway study.

### Data Collection and Outcomes

At each observation, participants received a full visual-tactile oral examination. Our primary outcomes were the incidence and prevalence of dental caries. Caries diagnosis was conducted following the International Caries Detection and Assessment System (ICDAS) adapted criteria for epidemiology and clinical research settings ^22^. Each tooth surface was assessed as being either intact/sound, sealed, restored, decayed, or arrested. Screening criteria considered lesions scored as a 5 (distinct cavity with visible dentin) or 6 (extensive, more than half the surface, distinct cavity with visible dentin) on the ICDAS scale as decay.

Every tooth and tooth surface was inspected for evidence of untreated decay. Any instance of decay was considered to be treatment failure, regardless of how many surfaces or teeth were affected. Additionally, any clinical presentation of a filling (e.g., amalgam, composite, stainless steel crown) on a tooth that previously was recorded as sound was similarly considered to be failure as it may be indicative of disease incidence in the time since the preceding observation.

Data were recorded using Electronic Health Record software designed for CariedAway (New England Software Systems, Boston, MA) and securely uploaded to a 128-bit encryption repository at the conclusion of each observation. Data were then processed at the Biostatistics and Epidemiology Data Analytics Center at Boston University, including checks for successful data transmission, congruence of unique patient identifiers with recorded values, and review for any out of range indicators for date of birth, school grade, presence/absence and number/type of teeth, and logical consistencies (e.g., change from decayed to sound). Any instances of errors upon review were then verified with individual performance sites (schools). Following quality control and assurance, data were then transmitted to New York University and locally stored on a secure server.

### Randomization

Schools were block-randomized to either the experimental or active control arm using a random number generator performed by RRR and verified by TBG.

### Blinding

Participants were blinded to their treatment assignments. However, due to the staining effect of silver diamine fluoride when applied to porous structure, patients would be able to derive their groups. Clinicians and examiners were not blinded as the procedures differed for each treatment, however clinicians were not able to discern who treated each participant at prior study observations.

### Impact of COVID-19

The original protocol for CariedAway included biannual data collection. However, due to the impact of the SARS-CoV-2 virus, schools in New York City were closed to healthcare providers from March 2020 through September 2021. Thus, the time elapsed between the observations corresponding to this period was approximately two years. Upon reopening of schools to healthcare services, the original schedule was resumed.

### Power

Sample size calculations for the longitudinal prevalence of caries for the CariedAway study were previously reported ^16^ assuming six observational periods, power of 0.80, a two-sided type I error rate of 5%, a repeated measures correlation of 0.5, and a per-visit attrition rate of 20%. Estimates also assumed a minimally detectable effect size of 0.25 and an intraclass correlation coefficient of 0.10, yielding a sample size of 12,874. However, at the completion of the trial the actual intraclass correlation coefficient across individual study observations ranged from 0.013 (prevalence) to 0.015 (incidence). As a result, the final participant enrollment was sufficient for power requirements.

### Statistical Analysis

We first organized participants by observation and computed individual descriptive statistics for sociodemographic and clinical variables for each study arm. At each observation, the proportion of participants in treatment groups with new caries or newly observed fillings was determined and bootstrapped confidence intervals for the difference were computed to account for any clustering effect of schools. The intraclass correlation coefficient for clinical outcomes was estimated using intercept-only mixed effects models.

We assessed longitudinal noninferiority using mixed effects logistic regression models, where the outcome was the presence or absence of any new caries at each observation. Models included random intercepts for individual participants and school. Our noninferiority margin of 10% was previously selected based on historical evidence and clinical judgement as to what would be an acceptable difference in efficacy for the prevention of dental caries ^23,24^. We converted the margin to the odds ratio scale by taking the average of the success proportion in the active control arm and determining the equivalent margin, yielding an ο_OR_ of 0.63 ^25^. We first tested noninferiority at any measurement period by including an interaction between treatment and time, followed by a model with no interaction to assess non-inferiority marginally. Comparisons to ο_OR_ were made using a (1-2α) confidence interval for the effect of treatment ^26^. Models adjusted for baseline decay, race/ethnicity, evidence of dental care received prior to study enrollment, and sex.

We calculated the incidence rate for the total number of individual teeth that developed caries (in tooth-years) and derived the rate ratio as the most efficient estimator due to the small degree of intra-cluster correlation in responses ^27^. We then modeled the per-person number of caries present at each observation using multilevel mixed-effects negative binomial regression. Pre-specified subgroup analyses for the effect of treatment over time and by the presence or absence of caries at baseline were performed. Covariates for incidence models were the same as those of prevalence.

Finally, we performed a series of supplementary analyses. To account for possible bias due to interval and right-censored observations, we analyzed cares incidence using Cox proportional hazards regression with nonparametric maximum likelihood estimation ^28^. We then assessed whether initial treatment by either a dental hygienist or registered nurse affected caries prevalence in the experimental group. Finally, we restricted analysis of caries prevalence and incidence to the subset of participants who were enrolled and received their first examination and treatment in the semester prior to school shutdowns due to COVID-19. This latter approach avoids differential rates of follow-up between each observation for participants in the analytic set.

Statistical significance was determined at p < 0.05. Analysis was performed in R v4.0 and Stata v16.

## Results

A total of 7418 participants were enrolled in CariedAway across 48 schools (Figure 1). After randomization, there were 3739 (50.4%) participants in the experimental group and 3679 (49.6%) participants in the active control group (Table 1). There were 4100 participants who completed at least one follow-up observation (55%), consisting of 2063 (50.32%) in the experimental group and 2037 (49.68%) in the active control. The total study observation time was 4 years: 507 days from baseline to first follow-up, 300 from first to second, 195 from second to third, 169 from third to fourth, and 171 from fourth to fifth. As a result of the two-year hiatus of health services in schools due to COVID-19, any enrolled participants in the fourth or fifth grades had aged out of the study upon resumption of data collection. The overall prevalence of baseline untreated caries was 26.7%, or 27.17% (95% CI = 25.7, 28.6) for the experimental group and 26.2% (95% CI = 24.8, 27.6) for the active control group in the enrolled sample. Untreated decay prevalence was similarly 28.3% (95% CI = 26.4, 30.3) for retained experimental participants and 27.3% (95% CI = 25.4, 29.3) for retained active control participants. The average age at baseline for the full sample was 7.58 years (SD = 1.90) and was comprised of 46% male. Approximately 75% of study participants in both groups reported as either Hispanic/Latino or black race or ethnicity.

**Figure 1:**
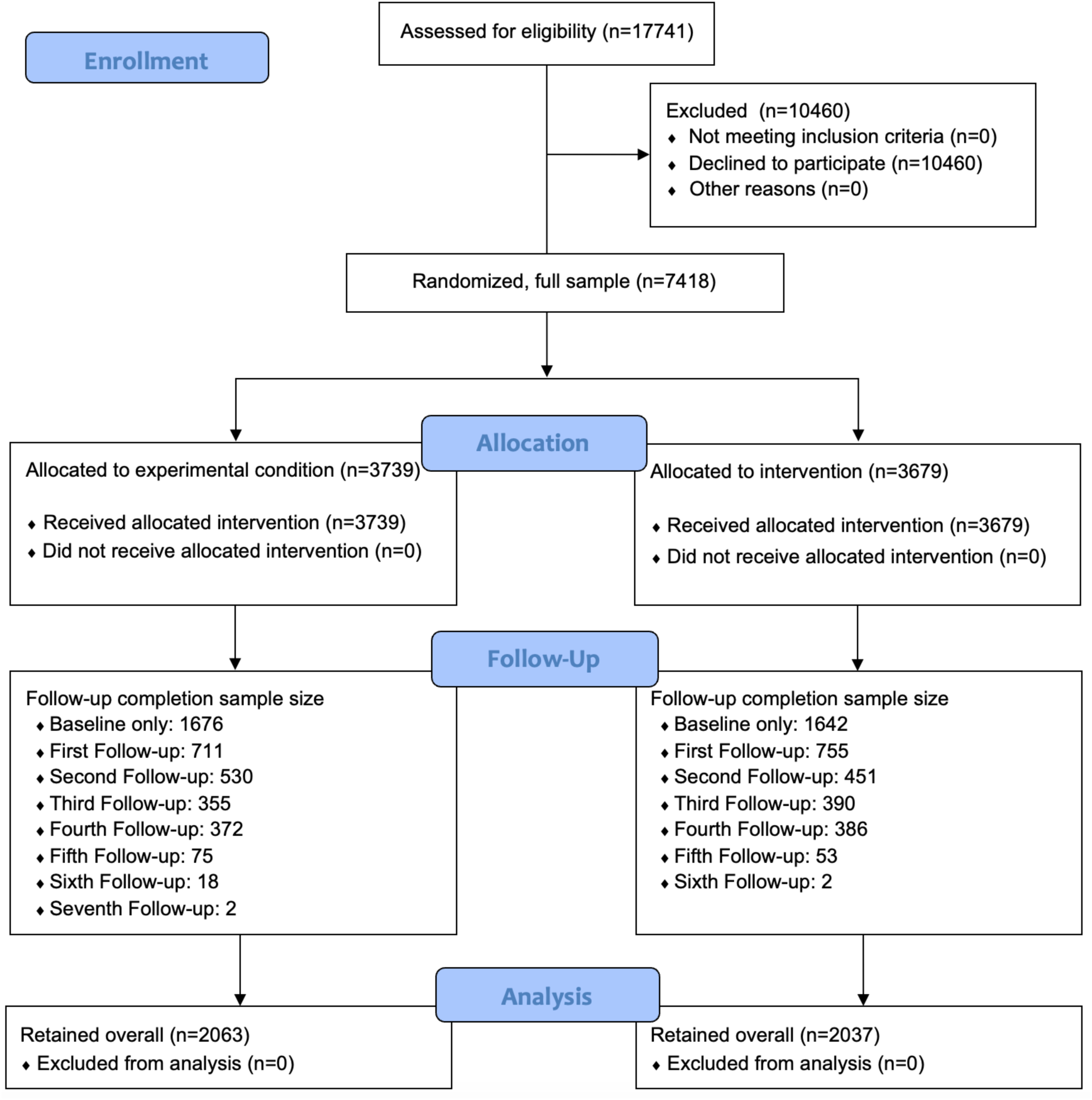
CONSORT Flow Diagram.

**Table 1:** Sample demographics overall and by treatment (enrolled and retained)

|  | Overall |  | Experimental |  | Active Control |  |
| --- | --- | --- | --- | --- | --- | --- |
|  | N | % | N | % | N | % |
| Participants (enrolled) | 7418 | 100 | 3739 | 50.4 | 3679 | 49.6 |
| Baseline decay | 1980 | 26.69 | 1016 | 27.17 | 964 | 26.2 |
| Sex (male) | 3412 | 46 | 1785 | 47.74 | 1627 | 44.24 |
| Race/Ethnicity |  |  |  |  |  |  |
| Hispanic/Latino | 3648 | 49.18 | 1766 | 47.23 | 1882 | 51.16 |
| Black | 1246 | 16.8 | 650 | 17.38 | 596 | 16.2 |
| White | 153 | 2.06 | 86 | 2.3 | 67 | 1.82 |
| Asian | 125 | 1.69 | 88 | 2.35 | 37 | 1.01 |
| More than one | 114 | 1.54 | 67 | 1.79 | 47 | 1.28 |
| Other | 90 | 1.21 | 56 | 1.5 | 34 | 0.92 |
| Unreported | 2042 | 27.53 | 1026 | 27.44 | 1016 | 27.62 |
| Age at baseline | 7.58 | 1.90 (SD) | 7.55 | 1.92 (SD) | 7.63 | 1.88 (SD) |
| Participants (retained) | 4100 | 100 | 2063 | 50.32 | 2037 | 49.68 |
| Baseline decay | 1140 | 27.80 | 584 | 28.31 | 556 | 27.30 |
| Sex (male) | 1872 | 45.67 | 975 | 47.26 | 897 | 44.06 |
| Race/Ethnicity |  |  |  |  |  |  |
| Hispanic/Latino | 2329 | 57.10 | 1155 | 55.99 | 1174 | 57.63 |
| Black | 794 | 19.47 | 416 | 20.16 | 378 | 18.56 |
| White | 86 | 2.11 | 56 | 2.71 | 30 | 1.47 |
| Asian | 78 | 1.91 | 59 | 2.86 | 19 | 0.93 |
| More than once | 62 | 1.52 | 40 | 1.94 | 22 | 1.08 |
| Other | 69 | 1.69 | 41 | 1.99 | 28 | 1.37 |
| Unreported | 682 | 16.63 | 296 | 14.35 | 386 | 18.95 |
| Age at baseline | 6.93 | 1.59 (SD) | 6.87 | 1.60 (SD) | 7.00 | 1.57 (SD) |

The prevalence of participants with no new caries or fillings at each observation (Table 2) show similar proportions in both groups, with differences in prevalence ranging from −.001 to 0.031 across study observations. For example, at the first follow-up observation recorded an average of 507 days post-baseline, the prevalence of participants with no untreated caries or newly observed fillings was 67% in the active control group and 64% in the experimental group, for a difference of 0.031. The bootstrapped 95% confidence interval for this difference was (−0.004, 0.067). For mixed-model analyses of caries prevalence over time (Table 3), the interaction effect between time and treatment was not significant, indicating that non-inferiority could be assessed marginally. Across both groups, the odds of untreated decay significantly decreased by approximately 21% at each observational visit (OR = 0.79, 95% CI = 0.75, 0.83). Expressed as the estimate for active control relative to experimental treatment for determining non-inferiority, the OR was 0.94 (95% CI = 0.80, 1.11 and 90% CI = 0.82, 1.08). The confidence interval for the marginal effect was outside the estimated 8_OR_ of 0.63 and non-inferior.

**Table 2:** Prevalence of participants without new caries or new fillings at each observation for active control (C) and experimental (T)

| Observation | Duration | C | T | C-T | 95% L | 95% U |
| --- | --- | --- | --- | --- | --- | --- |
| 2nd | 507 | 0.67 | 0.64 | 0.031 | -0.004 | 0.067 |
| 3rd | 300 | 0.69 | 0.69 | -0.001 | -0.042 | 0.039 |
| 4th | 195 | 0.7 | 0.7 | 0.003 | -0.047 | 0.051 |
| 5th | 169 | 0.76 | 0.75 | 0.01 | -0.029 | 0.059 |

*Notes: each upper bound of the 95% confidence interval is below the 10% non-inferiority threshold for the C-T difference. Any single instance of decay or new fillings not previously observed was considered treatment failure.*

**Table 3:**
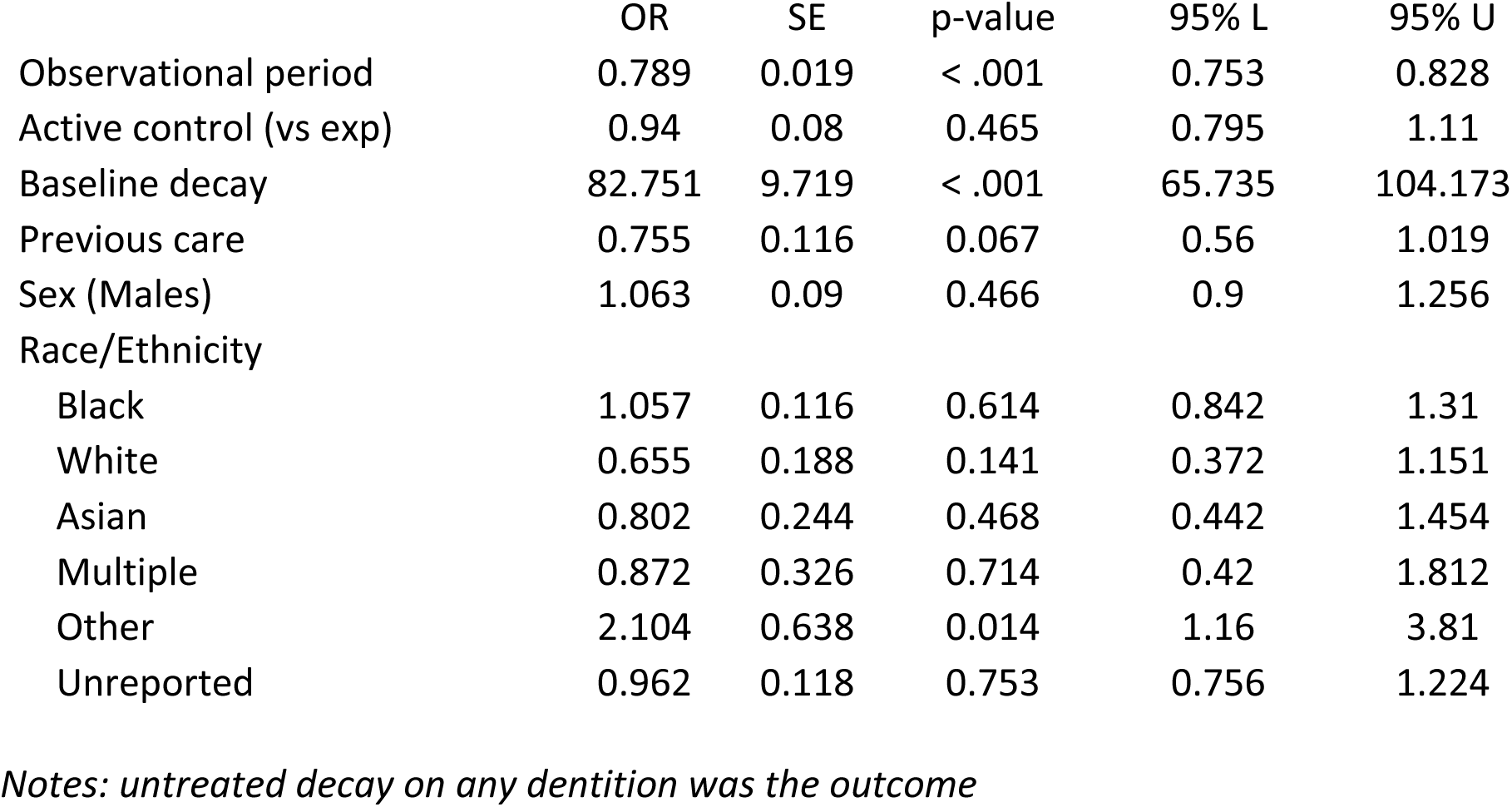
Longitudinal caries prevalence.

|  | OR | SE | p-value | 95% L | 95% U |
| --- | --- | --- | --- | --- | --- |
| Observational period | 0.789 | 0.019 | < .001 | 0.753 | 0.828 |
| Active control (vs exp) | 0.94 | 0.08 | 0.465 | 0.795 | 1.11 |
| Baseline decay | 82.751 | 9.719 | < .001 | 65.735 | 104.173 |
| Previous care | 0.755 | 0.116 | 0.067 | 0.56 | 1.019 |
| Sex (Males) | 1.063 | 0.09 | 0.466 | 0.9 | 1.256 |
| Race/Ethnicity |  |  |  |  |  |
| Black | 1.057 | 0.116 | 0.614 | 0.842 | 1.31 |
| White | 0.655 | 0.188 | 0.141 | 0.372 | 1.151 |
| Asian | 0.802 | 0.244 | 0.468 | 0.442 | 1.454 |
| Multiple | 0.872 | 0.326 | 0.714 | 0.42 | 1.812 |
| Other | 2.104 | 0.638 | 0.014 | 1.16 | 3.81 |
| Unreported | 0.962 | 0.118 | 0.753 | 0.756 | 1.224 |

*Notes: untreated decay on any dentition was the outcome*

For newly observed caries across the full study duration (Table 4), the crude incidence rate in the experimental group was 10.2 caries per 1,000 tooth-years. The rate in the active control was 9.8 caries per 1,000 tooth-years, for a rate ratio of 1.046 (95% CI = 0.973, 1.12) and a preventive fraction of 0.023. From adjusted models for longitudinal caries incidence (Table 5), the overall risk rate over time reduced (IRR = 0.83, 95% CI = 0.81, 0.85) with each observation. The risk comparing participants in the dental sealants with ART group to those in the SDF group was 0.924 (95% CI = 0.825, 1.035). There were no significant interactions between treatment and time and treatment and baseline decay status.

**Table 4:** Incidence rate of dental caries for experimental and active control.

|  | Experimental | Active Control | Total |
| --- | --- | --- | --- |
| Caries | 1625 | 1433 | 3058 |
| Tooth-years | 157979 | 145653 | 303632 |
| Incidence Rate | 0.0103 | 0.0098 | 0.0101 |
|  | Estimate | 95% CI |  |
| Incidence rate difference | 0.0004 | -0.0003, 0.0012 |  |
| Incidence rate ratio | 1.046 | 0.973, 1.123 |  |

**Table 5:** Longitudinal caries incidence.

|  | IRR | SE | p-value | 95% L | 95% U |
| --- | --- | --- | --- | --- | --- |
| Observational period | 0.828 | 0.009 | < .001 | 0.810 | 0.846 |
| Active control (vs exp) | 0.924 | 0.053 | 0.172 | 0.825 | 1.035 |
| Baseline decay | 17.277 | 1.018 | < .001 | 15.393 | 19.391 |
| Previous care | 0.758 | 0.076 | 0.005 | 0.624 | 0.922 |
| Sex (male) | 1.050 | 0.055 | 0.349 | 0.948 | 1.164 |
| Race/Ethnicity |  |  |  |  |  |
| Black | 0.987 | 0.068 | 0.854 | 0.863 | 1.130 |
| White | 0.721 | 0.133 | 0.076 | 0.503 | 1.035 |
| Asian | 0.907 | 0.170 | 0.602 | 0.627 | 1.310 |
| Multiple | 0.932 | 0.208 | 0.752 | 0.602 | 1.442 |
| Other | 1.742 | 0.304 | 0.001 | 1.237 | 2.452 |
| Unreported | 0.931 | 0.071 | 0.351 | 0.801 | 1.082 |

*Notes: total number of teeth at each observation with untreated dental caries was the outcome*

In supplementary analyses, following adjustment for baseline decay, receipt of prior preventive care, sex, and race/ethnicity, the hazard ratio comparing the active control to experimental for time to first observed carious lesion was 0.91 (95% CI = −.826, 1.08, table not shown). Post-estimation inspection of Turnbull’s nonparametric and Cox predicted survival curves indicated that the data did not violate the proportional-hazards assumption. Additionally, there were no significant differences in caries prevalence in children treated with SDF by registered nurses compared to dental hygienists (OR = 0.89, 95% CI = 0.67, 1.19, table not shown), and the provider effect did not significantly change over time (OR = 1.00, 95% CI = −.85, 1.17).

For the sub-sample, 4718 CariedAway participants were enrolled, randomized, and treated in the semester prior to school closures resulting from COVID-19 (September 2019 through March 2020), of which 2998 were viable for follow-up after pandemic restrictions were lifted (Supplementary Figure 1). At the completion of the trial, follow-up data were obtained for 1831 participants. A total of 380 (12.7%) participants successfully completed one follow-up observation, 416 (13.9%) participants completed two, 430 (14.3%) completed three, 563 (18.8%) completed four, and 42 (1.4%) completed five. The corresponding duration in days for each follow-up observation for the sub-sample was 768, 193, 187, 161, and 163, for a total of 1472 days or 4.03 years. Compared to the full sample, the longer duration from baseline to first follow-up was due to study suspension in response to COVID-19. Approximately 29% of children in the sub-sample had untreated decay at baseline (Supplementary Table 1) and the average age was 6.63 (SD=1.24). Results for longitudinal analyses for caries prevalence (Supplementary Table 2) and incidence (Supplementary Table 3) were similar to that of full-sample analyses. The odds ratio comparing active control to SDF-treated participants for caries prevalence was 1.04 (95% CI = 0.82, 1.31) and non-inferior. Similarly, the incident rate ratio for total number of decayed teeth was 0.995 (95% CI = 0.851, 1.163). Similarly, the odds of decay prevalence and incidence significantly decreased with each observational period.

## Discussion

School sealant programs have demonstrated effectiveness in reducing the risk of dental caries ^29^, yet are underused due to the burdensome costs of care ^9^. Many children subsequently continue to suffer from untreated disease, which can lead to systemic infection and negatively affect child development ^30^. Our overall results show that application of silver diamine fluoride with fluoride varnish was non-inferior compared to dental sealants, fluoride varnish, and ART in the longitudinal prevalence of caries when used in a school program. We conclude that silver diamine fluoride is an effective alternative for community-based prevention that can address these existing barriers.

Although silver diamine fluoride is primarily used as a caries arresting agent, it is also effective in the prevention of new caries ^31,32^. There is a reduced risk of new caries on surrounding sound dentition when existing lesions are treated ^33^ and SDF is more effective than fluoride varnish in preventing new caries in early childhood ^34^. However, prior short term comparative assessments of SDF yields conflicting results on its superiority relative to glass ionomer sealants and atraumatic restorations. ^35,36^. These previous trials were also restricted to either 12 or 24 months of observation, and little long-term evidence exists ^10,24^.

In addition to clinical effectiveness, the simplicity and financial implications of a school-based silver diamine fluoride program can result in considerable cost savings to the public. A review of existing SDF treatment protocols identified application times as low as 10 seconds per tooth ^12^, suggesting that more children can be treated in less time. Use of SDF as a caries management strategy also reduces Medicaid program expenditures ^37^, is the most cost-effective option in populations with a high risk of dental caries ^38^, and is more cost-efficient compared to ART ^13^, although potential restrictions from Medicaid reimbursement may persist ^37^.

In 2022, the American Medical Association approved a category III Current Procedural Terminology code authorizing non-dental healthcare professionals to administer SDF, supporting an expansion of services into alternative settings and providers. A secondary objective of CariedAway was to assess the effectiveness of nurses in the use of SDF, and we previously showed that registered nurses were non-inferior compared to dental hygienists in the prevention of dental caries ^20^. Similarly, our presented results for the prevalence and incidence of dental caries over four years included participants treated by either hygienists or nurses. Within the SDF arm of the CariedAway trial, approximately 20.5% of all baseline participants and 13.5% of all individual participant encounters were treated by registered nurses. Implementation of school sealant programs previously found greater student participation when school nurses partner with hygienists in the delivery of care ^39^, but our results empower nurses themselves as primary providers for caries prevention. With over 132,000 school nurses estimated to be currently working in the United States ^40^ and the growing involvement of school nurses in oral health promotion and prevention ^41^, these findings can expand the scope of practice for both school nurses and nurses in family practices, dramatically increasing access to care.

Approximately 1 in 4 of children participating in CariedAway had untreated disease at baseline (1 in 3 for the COVID-19 sample), and we previously showed that only 11% had pre-existing sealants at their time of enrollment ^42^. Following treatment, the overall odds of dental caries decreased by approximately 20% in both study arms. The risk of incident dental caries was nearly identical in both treatment groups, resulting in a very small preventive fraction between the included interventions. This was not unexpected given the non-inferiority design. Similarly, the data indicate no significant differences across treatment in the risk of first caries eruption or when modeling the total number of new dental caries experienced overall, nor is there sufficient evidence to indicate whether there are differences in treatment effect over time or based on the presence of disease at baseline. Prior research with dental sealants estimates a 50% preventive fraction compared to placebo; comparisons of children with and without dental sealants concludes that the prevention of over three million cavities would be attributable to sealants ^43^. The similarity in observed incidence from CariedAway may support a similar conclusion for the application of silver diamine fluoride.

While the American Dental Association ^44^ and the American Academy of Pediatric Dentistry ^45^ include silver diamine fluoride in their clinical recommendations for caries management, known complications with SDF application include potential oral soft tissue irritation, temporary staining of the oral mucosa, and permanent staining of porous tooth structure such as dental caries or hypomineralization ^31^. Despite thousands of SDF applications in CariedAway, we encountered no adverse events including mild reports (no intervention required; no impact on activities of daily living [ADL]), moderate (minimal, local, or non-invasive intervention indicated, moderate impact on ADL), or severe (significant symptoms requiring invasive intervention; subject seeks major medical attention, needs major assistance with ADL) and received only one complaint regarding staining, which pertained to superficial skin staining from accidental spillage that was mistaken for bruising. Our prior findings from CariedAway similarly did not indicate a negative impact of SDF therapy on oral health-related quality of life, which included measures for aesthetic perceptions of the oral cavity ^18^. Other research concludes that a high proportion of parents of children treated with silver diamine fluoride remain satisfied with their child’s dental appearance ^46,47^, that aesthetic concerns are mitigated with posterior application ^48^, and that no differences were found in adverse events or aesthetic perceptions when comparing children treated with SDF or ART ^49^. Our results suggest that use of SDF in a school-based program is well tolerated by both children and their caregivers.

A total of 167 primary schools in New York City met eligibility criteria for the CariedAway trial, serving over 87,000 students consisting of approximately 69% Hispanic/Latino, 95% at or below 135% of the federal poverty level, and 60% participating in Medicaid. All schools were approached for participation in the study. Of the 48 schools electing to participate in CariedAway across thirteen districts, 64.7% of the student population identified as Hispanic and 26% as black, and 86% reported living below 135% of the federal poverty level. At the student level, there were 1,047,895 students in the NYC school system for the 2022-2023 academic year, consisting of 72.8% economically disadvantaged, 41.1% Hispanic, 23.7% black, 16.5% Asian, and 14.7% white. Compared to these proportions, the CariedAway participants slightly overrepresented Hispanic children and underrepresented black children, however 27% of participants did not report their race/ethnicity.

As a pragmatic trial, there are concerns regarding subject attrition and the potential bias from external care. Our analysis used all available observations for study participants and also considered a subset of participants that had equal rates of follow-up to address the disruption in study activities due to the effects of COVID-19. Although prior results from CariedAway concluded that a single application of SDF and varnish was non-inferior to sealants/ART for caries arrest after two years, analysis was restricted to children who did not have dental caries at baseline ^17^. The presented findings assessed non-inferiority inclusive of both children who did or did not begin the study with active untreated decay, which has previously not been reported. The inclusion of participants with untreated dental caries at study start results in a higher risk of subsequent disease development, and addresses concerns that dental caries incidence rates in children who present with no evidence of decay may be driven largely by better oral hygiene behavior, dietary intake, or access to routine dental cleanings, instead of the included interventions. Additionally, our assessment of dental caries incidence included not only active decay but any evidence of decay that was treated by an external clinician. We also included multiple approaches for prevention, including any incidence of decay, overall prevalence, time to first eruption, and estimates at both the tooth and person levels. Our analysis further adjusted for censoring, a common issue in the assessment of school-based programs. While attrition is a clear weakness, the pragmatic nature of the trial is also its strength. Our results reflect the practical, real-world impact of a school-based oral health model that utilizes silver diamine fluoride for long-term caries management.

In conclusion, untreated dental caries has maintained a sequential, thirty year position at the top of the global disease prevalence list in low, middle, and high income countries ^50^. In response, the World Health Organization included both silver diamine fluoride and glass ionomer on its inaugural list of essential dental medicines for a basic health-care system ^4^. Our results provide quantitative, longitudinal evidence on the comparative impact of these essential medicines, with particular application for community-based prevention. The simplicity, cost-efficiency, and versatility of silver diamine fluoride can be used by clinicians, practices, communities, and countries in the global pursuit of the WHO Global Oral Health Action Plan ^3^.

## Author Contribution

RRR and RN conceived of the CariedAway study, obtained funding, and were the principal investigators. RRR wrote the manuscript and performed analyses. TBG directed clinical operations, collected data, and managed study clinical teams. All authors critically revised and approved the manuscript.

## Data Availability

Data
Data available: Yes
Data types: Data dictionary
How to access data: Data dictionaries will be available to interested researchers upon request to the authors
When available: beginning date: 09-01-2023
Supporting Documents
Document types: Informed consent form
How to access documents: Informed consent forms will be available to interested researchers upon request to the authors
When available: beginning date: 09-01-2023
Additional Information
Who can access the data: Interested researchers upon request to the authors
Types of analyses: For any purpose
Mechanisms of data availability: After approval of a proposal and a signed data access agreement.

---

**Supplementary Figure 1:**
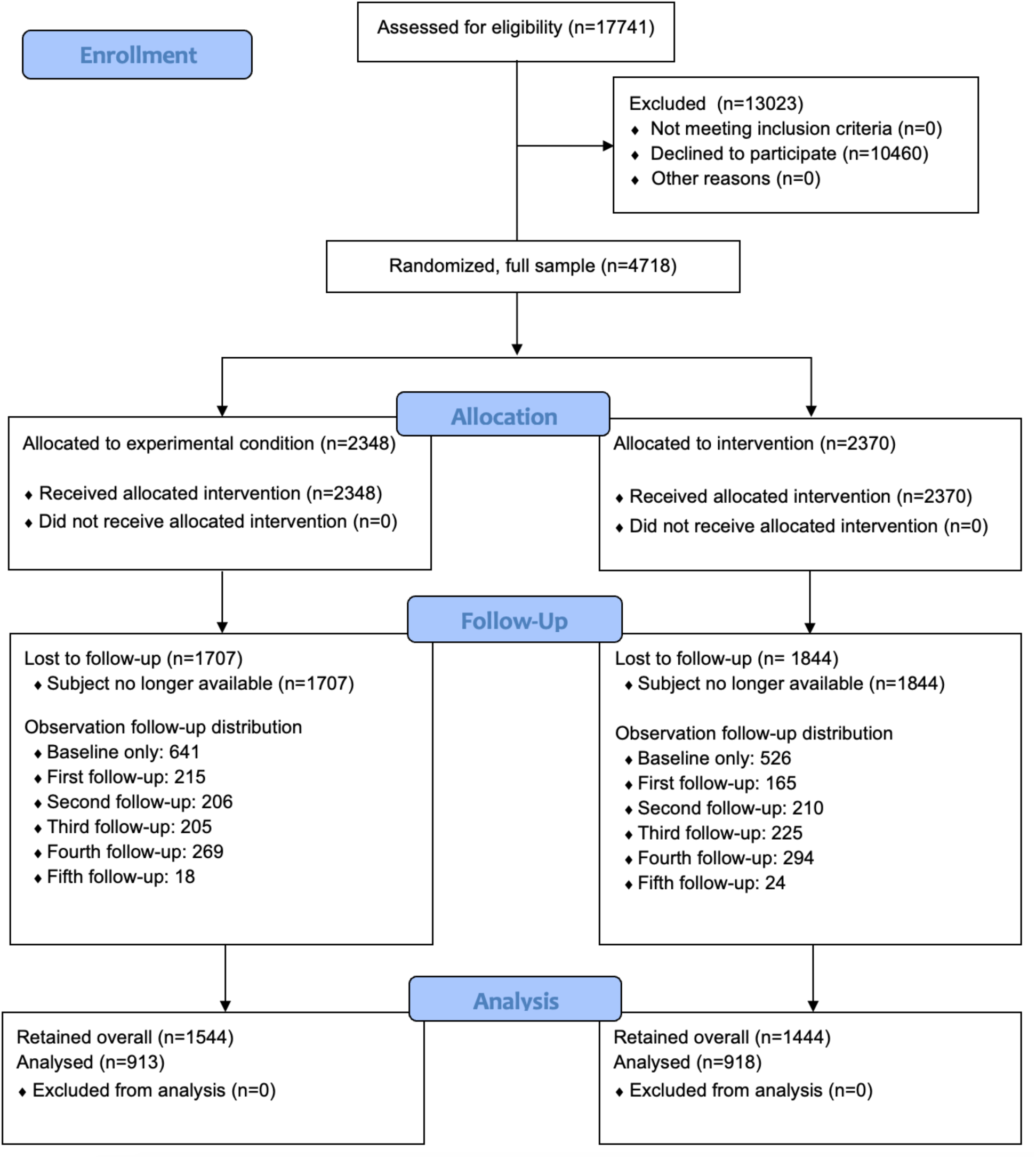
CONSORT diagram, COVID-19 sample.

**Supplementary Table 1:** Sample demographics overall and by treatment (COVID-19 sample)

|  | Overall |  | Experimental |  | Active Control |  |
| --- | --- | --- | --- | --- | --- | --- |
|  | N | % | N | % | N | % |
| Participants | 2998 | 100 | 1554 | 51.83 | 1444 | 48.17 |
| Baseline decay | 874 | 29.15 | 482 | 31.02 | 392 | 27.15 |
| Sex (male) | 1432 | 47.77 | 664 | 45.98 | 769 | 49.42 |
| Race/Ethnicity |  |  |  |  |  |  |
| Hispanic | 1631 | 54.4 | 810 | 52.12 | 821 | 56.86 |
| Black | 558 | 18.61 | 311 | 20.01 | 247 | 17.11 |
| White | 75 | 2.5 | 36 | 2.32 | 39 | 2.7 |
| Asian | 44 | 1.47 | 29 | 1.87 | 15 | 1.04 |
| More than one | 58 | 1.93 | 34 | 2.19 | 24 | 1.66 |
| Other | 41 | 1.37 | 24 | 1.54 | 17 | 1.18 |
| Unreported | 591 | 19.71 | 310 | 19.95 | 281 | 19.46 |
| Age at baseline | 6.63 | 1.24 (SD) | 6.62 | 1.25 (SD) | 6.65 | 1.24 (SD) |

*Notes: sample includes any subject that (1) was enrolled in the six-month data collection period prior to school shutdowns due to COVID-19 and (2) completed a follow-up observation upon resumption of study activities .*

**Supplementary Table 2:** Longitudinal caries prevalence (untreated caries), COVID-19 sample.

|  | OR | SE | p | 95% L | 95% U |
| --- | --- | --- | --- | --- | --- |
| Observational period | 0.789 | 0.025 | < .001 | 0.741 | 0.84 |
| Active control (vs exp) | 1.04 | 0.125 | 0.763 | 0.819 | 1.31 |
| Baseline decay | 46.57 | 7.18 | < .001 | 34.43 | 62.99 |
| Previous care | 0.744 | 0.152 | 0.148 | 0.499 | 1.11 |
| Sex (Males) | 1.04 | 0.125 | 0.751 | 0.821 | 1.31 |
| Race/Ethnicity |  |  |  |  |  |
| Black | 1.04 | 0.158 | 0.797 | 0.772 | 1.4 |
| White | 0.89 | 0.342 | 0.757 | 0.418 | 1.89 |
| Asian | 0.489 | 0.224 | 0.118 | 0.2 | 1.2 |
| Multiple | 0.663 | 0.363 | 0.454 | 0.227 | 1.94 |
| Other | 2.05 | 0.844 | 0.08 | 0.917 | 4.59 |
| Unreported | 1.07 | 0.205 | 0.723 | 0.735 | 1.56 |

**Supplementary Table 3:** Longitudinal caries incidence, COVID-19 sample.

|  | IRR | SE | p-value | 95% L | 95% U |
| --- | --- | --- | --- | --- | --- |
| Observational period | 0.816 | 0.013 | < .001 | 0.792 | 0.841 |
| Active control (vs exp) | 0.995 | 0.079 | 0.948 | 0.851 | 1.163 |
| Baseline decay | 12.678 | 0.106 | < .001 | 10.770 | 14.930 |
| Previous care | 0.776 | 0.103 | 0.056 | 0.598 | 1.001 |
| Sex (male) | 1.030 | 0.078 | 0.651 | 0.893 | 1.120 |
| Race/Ethnicity |  |  |  |  |  |
| Black | 1.000 | 0.097 | 0.964 | 0.832 | 1.213 |
| White | 0.878 | 0.217 | 0.598 | 0.540 | 1.426 |
| Asian | 0.629 | 0.187 | 0.119 | 0.351 | 1.127 |
| Multiple | 0.728 | 0.244 | 0.345 | 0.377 | 1.406 |
| Other | 1.560 | 0.392 | 0.078 | 0.952 | 2.553 |
| Unreported | 0.981 | 0.117 | 0.873 | 0.776 | 1.240 |

